# Uncovering Social Media Influencers’ Health Posts: A Youth-Driven Mixed Method Study

**DOI:** 10.64898/2026.10.13.26365375

**Authors:** Gell Sascha, Engel Elena, Karsay Kathrin, Heiss Raffael

**Author notes:** Corresponding Author: Gell Sascha, Universitätsstraße 15, MCI | The Entrepreneurial School, 6020, Innsbruck, Austria. Elena Engel, Universitätsstraße 15, MCI | The Entrepreneurial School, 6020, Innsbruck, Austria, Kathrin Karsay, Kolingasse 14-16, University of Vienna, 1090 Wien, Austria, Raffael Heiss, Universitätsstraße 15, MCI | The Entrepreneurial School, 6020, Innsbruck, Austria.

## Abstract

Social media influencers serve as a key source of health information for adolescents but often lack expertise, have commercial motives, and may spread misleading or harmful content. This study examines the health-related content shared by influencers, the prevalence of advertising, and how adolescents evaluate potential benefits and risks. Using a mixed-methods approach, we employed a quantitative descriptive method involving citizen science (Study 1) and a qualitative content analysis (Study 2). In Study 1, 336 adolescents from 16 Austrian schools documented and categorized 2,147 posts from 255 influencers. The most common topics included sports and fitness (50.5%), beauty ideals (29.6%), food and nutrition (27.6%), and cooking (19.1%). Although advertisements appeared in 41.5% of posts, adolescents rated 60.4% of them as beneficial and only 4.5% as harmful. The qualitative analysis of a subsample of the 90 most prominent posts showed that adolescents appreciated motivational aspects (e.g., healthy eating) but often overlooked problematic elements such as misleading claims, sexualized body ideals, and the linking of health messages to commercial products. Enhancing persuasion knowledge and critical thinking through education, along with stricter regulation of harmful products, is crucial to safeguarding youth online.

## 1. Introduction

Social media influencers have become an important source of young people’s health information (Adeane and Stasiak, 2024; boyd, 2019; Nygård and Lindfors, 2025). They are defined as “individuals who amass large followings on social media and exert significant influence over their audience through engaging content” (Engel et al., 2024a). In a recent representative survey from Austria, 15-to-25-year-olds report that influencers cover more than half of the content they see on social media, and 83% say that they encounter health-related content from influencers at least sometimes (Engel et al., 2024b).

These numbers are critical given that influencers are perceived as trustworthy sources by adolescents, offering alternative information to traditional sources such as physicians or health organizations (Kostygina, 2020; Lim et al., 2022). While some influencers may promote healthy lifestyles and provide valuable health information, researchers have also identified significant challenges. For instance, influencers might lack formal training, could be biased due to industry interests, or share misleading health information and advice (Raaper et al., 2024; Suarez-Lledo and Alvarez-Galvez, 2021; Wang and Togher, 2024).

Despite these pressing challenges, there is limited understanding of how adolescents – the main audience of influencer communication – engage with these new agents and their content (Engel et al., 2024a; Fu & Cook, 2020; Powell and Pring, 2024). One reason for this gap is that social media environments are considered “black boxes” to researchers, concealing information about personal preferences, click behavior, and algorithmic decisions from the public (Krause et al., 2025). As a result, what young people encounter is often not directly accessible to researchers.

While traditional scientific methods are not well equipped to address this challenge, citizen science offers a promising way to approach personalized social media environments by leveraging crowdsourcing processes (Heiss and Matthes, 2017; Qutteina et al., 2019). In citizen youth science, young volunteers take on the roles of professional researchers. Their roles can vary from handling one aspect within the research process (e.g., data collection) to participating in all steps of the research, including design, data analysis, and interpretation. In addition, engaging laypeople as citizen scientists in medical and public health research provides several benefits, including empowering affected populations, educational impacts, inclusive policymaking, and improved community health (Den Broeder et al., 2018; Fiske et al., 2019; Psaroudakis et al., 2020; Bonney et al., 2009).

### Study 1

To address these challenges the first study focused on a citizen science project where adolescents documented and categorized influencer profiles and health-related posts. To address the research gaps regarding adolescents’ exposure to and perception of health content from influencers we examined the following exploratory research questions:

**RQ1:** What are the characteristics of influencers posting about health, and what health content is disseminated by them?

**RQ2:** To what extent does advertisement permeate health content?

**RQ3:** How do adolescents evaluate the content disseminated by these influencers?

### Study 2

Study 2 was directly informed by the findings of the citizen science project (Study 1), which provided valuable insights into how citizen youth scientists (CYS) evaluated influencer-shared health content and identified the prevalence of advertisements within these posts. In Study 1, the health content disseminated by influencers was predominantly rated as beneficial by the CYS, despite the notable presence of a high density of advertisements.

This observation highlighted a potential discrepancy between the perceived health benefits of the content and the commercial motives embedded within it and raised critical questions about adolescents’ ability to critically evaluate health-related content. This prompted the research team to conduct Study 2, focusing on an in-depth qualitative analysis of the most prominent posts documented by the CYS. The fourth research question was developed to assess differences in the evaluation of posts between the CYS and the research team, aimed to uncover misconceptions and provide deeper insights into how adolescents interpret health-related social media posts:

**RQ4:** How do students’ assessments of health influencers’ posts, in terms of health impact and advertisement density, align with those of the research team?

## 2. Theoretical Background

The rise of social media influencers as health communicators is rooted in a shifting media landscape, where traditional gatekeepers – such as physicians, teachers, and public health institutions – now compete with decentralized, peer-like communicators on social media (Heiss et al., 2025; Lim et al., 2022). This transformation is particularly evident among adolescents, who perceive influencers as relatable and accessible sources that align with their digital habits and developmental needs (Engel et al., 2024a; Fu and Cook, 2020). In addition, algorithmic curation likely amplifies this dynamic by prioritizing influencer content targeted at young users (Cotter, 2019).

From a social learning perspective, influencers can serve as role models whose behaviors, values, and health-related norms are observed and potentially imitated by adolescents (Bandura, 2002; Engel et al., 2024a). They are also perceived as trustworthy sources, in part because of their ability to build parasocial relationships with young people – attachments formed through repeated exposure to an influencer’s content that resemble actual, but one-sided, social relationships in structure and psychological function (Giles, 2002; Yuan & Lou, 2020).

At the same time, the quality of health-related content shared by influencers varies widely and is often designed to be entertaining and emotionally engaging (Engel et al., 2024a; Powell & Pring, 2024). Initial content analyses indicate that such content is frequently biased due to commercial interests. For example, Nickel and colleagues (2024) found that influencers promote medical self-tests by emphasizing potential benefits while downplaying risks, thereby increasing the likelihood of overdiagnosis. Other studies have shown that influencers often endorse unhealthy food products (Qutteina et al., 2019), and e-cigarette use (Vassey et al., 2020; Vassey et al., 2022), but also communicate unrealistic and idealized body images (Ahrens et al., 2022).

As a result, health advice conveyed by influencers is often intertwined with commercial motives and embedded within advertising messages (Engel et al., 2024a). Frequently, such health content appears in promotional or lifestyle-oriented formats – designed to attract attention and persuade users – but these formats often lack informational accuracy or transparency (Borchers, 2022; Pilgrim & Bohnet-Joschko, 2019; van der Bend et al., 2022). To navigate this environment, adolescents need a broad set of critical evaluation skills, including information literacy, persuasion literacy, and health literacy (Heiss et al., 2024; Naderer, 2021; Nutbeam, 2021). These competencies help them assess the credibility of content, recognize persuasive intent, and identify misleading or biased claims.

Even though influencers have emerged as prominent and appealing sources of health information, there is still a lack of studies that use participatory approaches (such as citizen science) to systematically examine the health-related topics and commercial products that adolescents are exposed to when engaging with influencer content. The first study addresses these gaps by analyzing influencer content sampled by adolescents themselves and by directly involving them in the evaluation process. The second study aims to achieve a comprehensive understanding of the content that youth are exposed to through qualitative content analysis. Further, it seeks to examine how young people evaluate such health-related content in terms of its potential impact on health.

## 3. Method

### 3.1 Study 1: Citizen Science

The citizen science study employed a contributive approach in which adolescents acted as citizen youth scientists (CYS), participating in data collection and categorization (Den Broeder et al., 2018). The study received ethical approval by the IRB of the MCI | The Entrepreneurial School® (reference number: 20230312). Participants were recruited via existing school networks and the “Citizen Science Award” initiative – an annual project initiated by the Austrian Ministry of Education, Science and Research. The goal of the initiative is to enhance exchange between schools and academic partners and motivate young people to engage in research.

Sixteen schools in Austria, comprising 19 classes with 336 students (aged 12–19 years; 210 girls, 115 boys, and one non-binary student), participated in the citizen science study. The participating students ranged from seventh to eleventh grade, with seven schools located in rural and nine in urban areas.

The research team provided online information materials and a video tutorial on using the data collection tool to brief teachers and students about the project, research processes, methods, data collection, and categorization.

### 3.2 Quantitative Data Collection

The research team designed an online data collection tool, implemented as a Google Sheets spreadsheet, for adolescents to document influencer profiles and health-related social media posts. We pretested the tool with a middle school class of 18 students and two teachers. It was accessible on PCs, tablets, and smartphones via Google Tabs. The tool consisted of two sections: one for profile-level data of influencers (e.g., profile name, platforms, age), another one for post-level data (e.g., health topic, impact, advertisements) (Supplementary eTable 1). Each participating class selected a week between April and May 2023, during which CYS collected data. The classes identified and decided which influencers they considered the most important after consulting their schoolmates in a plenary session. They then documented at least five health-related and publicly available social media posts per chosen influencer using the data collection tool. However, most classes submitted more posts than the minimum required. Data collection focused on Instagram, TikTok, and YouTube since these were the most popular social media platforms among adolescents in Austria at the time (Saferinternet, 2023).

### 3.3 Quantitative-Descriptive Analysis

The data were analyzed using descriptive statistics and qualitative analysis. In the first step, the researchers examined the profile-level data for the characteristics of influencers (e.g., age, gender, platform), followed by the analysis of post-level data (e.g., health topic, health impact, advertisements). It is important to note that some questions allowed for multiple classifications, including used platforms, languages, and main topics on the profile level and post formats, health topics, advertisement types, and product types on the post level (Supplementary eTable 2, eTable 3).

### 3.4 Study 2: Qualitative Content Analysis

In Study 2, we conducted a qualitative content analysis to gain a more comprehensive understanding of the posts’ content and how the young students perceived them in terms of health impact compared to the researchers. To this end, we analyzed a subset of 90 posts, consisting of the ten posts with the highest engagement for each of the nine health topics addressed during the citizen science study (see Supplementary eTable 1). This additional qualitative analysis provided deeper insight into how the CYS categorized the health impacts of the posts, the extent to which they were able to identify disclosed and undisclosed advertisements, and the presence of potentially harmful or misleading content youth are exposed to.

### 3.5 In-depth Analysis

Three researchers independently reviewed and summarized the content of the subsample of 90 posts (see Table 1). The unit of analysis included either a single picture with a caption, an array of pictures with a caption, or a video. The researchers first examined the posts for disclosed and undisclosed advertisements and categorized them according to the data collection tool used by the CYS (i.e., addictive substances, sports equipment, food and nutrition, beauty and lifestyle, technical devices, apps, workshops or workout programs, and other products).

**Table 1.** Selected influencer posts inaccurately classified as ‘beneficial’ by adolescents.

| Topic | Influencer | Post description | Advertisement | Harmful content |
| --- | --- | --- | --- | --- |
| Beauty Ideal | YouTube:<br>Cloe Ting<br>Follower:<br>24.500.000 | The video shows the influencer doing a workout while promising that following this routine for two weeks will lead to considerable weight loss. | Clothing and workouts series of own product line | Focusing on weight loss as primary exercise goal, and promoting a sexualized and for many unrealistic body ideals. |
| Food Supplement | Instagram:<br>alex_eubank31<br>Follower:<br>2.300.000 | Several pictures of an influencer showing his body transformation while promoting several products. | Food supplements and energy drink (Alpha Lion, Celsius)<br>Ready-to-eat meals (FlexPro meals) | Promotion of sexualized and questionable beauty ideal.<br>Promotion of processed foods and food supplements. |
| Cooking | Instagram:<br>leanadeep<br>Follower:<br>4.700.000 | The influencer is portrayed in a video preparing breakfast, lunch, and dinner. Recipes are provided in the caption. The preparation is often interrupted by the consumption of protein drinks and training sequences. | Food Supplements of own product line | Suggesting that healthy nutrition and workouts need to be accompanied by the consumption of promoted food supplements. |
| Sports, Training & Fitness | YouTube:<br>Lilly Sabri<br>Follower:<br>4.500.000 | The video depicts the influencer working out while promoting her own app. Promises are made verbally and in the caption to lose belly fat within seven days of following her routine. The caption advertises her cookbook, promising to prevent inflammation and strengthen the immune system. | App and cookbook of own product line<br>Food supplements (Optimum Nutrition) | Promotion of a sexualized and questionable beauty ideal. Focus on weight loss without addressing other health benefits.<br>Misleading promises of visible physical changes and health benefits. Promotion of food supplements. |

In the next step, the researchers screened the posts for potentially harmful or misleading health content. This included the advertisement and/or promotion of addictive substances such as alcohol, illicit drugs, cannabis, or nicotine products (Vassey et al., 2020; Vassey et al., 2022); the promotion of potentially harmful beauty ideals, including thin, muscular, or “fit” body ideals and sexualized exposure (Thompson et al., 1999), which may contribute to body dissatisfaction and negative health outcomes such as eating disorders, depression, social anxiety, and suicidality (Fioravanti et al., 2022); the promotion of processed foods high in fat, salt, and sugar as well as sugar-sweetened beverages, which are associated with adverse health outcomes such as obesity, diabetes, and chronic diseases (Malik et al., 2006; Olsen & Heitmann, 2009; World Health Organization, 2023); and the promotion of untested food supplements that may contain unapproved pharmaceutical ingredients (Tucker et al., 2018) or contaminants such as pesticides, metals, or toxins (Costa et al., 2019).

Finally, the researchers compared their analyses with the coding of the CYS to identify potential misconceptions among adolescents. Intercoder reliability was established by comparing the three researchers’ codes. An agreement was reached for 84 posts (93.3%) and inconsistencies were discussed in the group and resolved collaboratively.

## 4. Quantitative Results (Study 1)

The participating adolescents documented 2147 posts from 255 influencer profiles. The profiles had a combined total of more than 1.6 billion followers distributed across Instagram, TikTok, and YouTube.

### 4.1 Platforms and Influencers

Most influencers (61.57%) appeared on all three platforms, while 11.37% of the identified influencers were present on only one platform. Since Instagram was the most popular platform among Austrian adolescents at the time, it is not surprising that 95.3% of the identified profiles were from Instagram. The profiles included of 5.5% micro-influencers (with less than 20,000 followers), 12.6% meso-influencers (20,000 to 100,000 followers), 16.5% macro-influencers (100,000 to 300,000 followers), and 67.5% celebrity-influencers (300,000+ followers). CYS identified more women (56%) than men (40%; 4% not identifiable), primarily within the age range of 19 to 39 years, with no influencer exceeding the age of 60. Most influencers came from German-speaking European countries (i.e., Austria, Germany, and Switzerland), accounting for 61.5%, followed by 22% from the United States, Canada, and Australia, 9.8% from non-German-speaking European countries, and 4.7% from other regions. (Supplementary eTable 2).

### 4.2 Post Characteristics

More than half (56.5%) of the 2147 posts were published on Instagram, 24.1% on TikTok, and 18.7% on YouTube. Posts primarily had an audio-visual format (73.8%); posts containing only pictures were the least utilized post format (32.5%). Post engagement ranged from six interactions (likes and comments) to 13,291,689 interactions. However, the mean engagement rate per post was 101,894 likes and comments (Supplementary eTable 3).

### 4.3 Health Content and Impact

The key health topics identified were sports and fitness (50.5%), beauty ideals (29.6%), food and nutrition (27.6%), and cooking (19.1%). Posts on the topics of mental health (12%), food supplements (7.5%), sexual and reproductive health (3.4%), and substance use (0.9%) were less frequent. Adolescents mostly viewed the posts (60.4%) as beneficial for their health, while in contrast, only a minority (4.5%) were seen as harmful (Supplementary eTable 3).

CYS documented 104 posts (5.5%) with specific illnesses, covering various health-related issues not specified in the data collection tool. Most notably, posts presented behavioral, mental, or neurodevelopmental disorders (e.g., depression, anxiety, eating disorders), skin-related health issues (e.g., cellulite, acne, eczema, lipoedema), diseases of the genitourinary system (e.g., infertility, endometriosis), infectious diseases like COVID-19, or endocrine, nutritional, or metabolic diseases.

**Figure 1.**
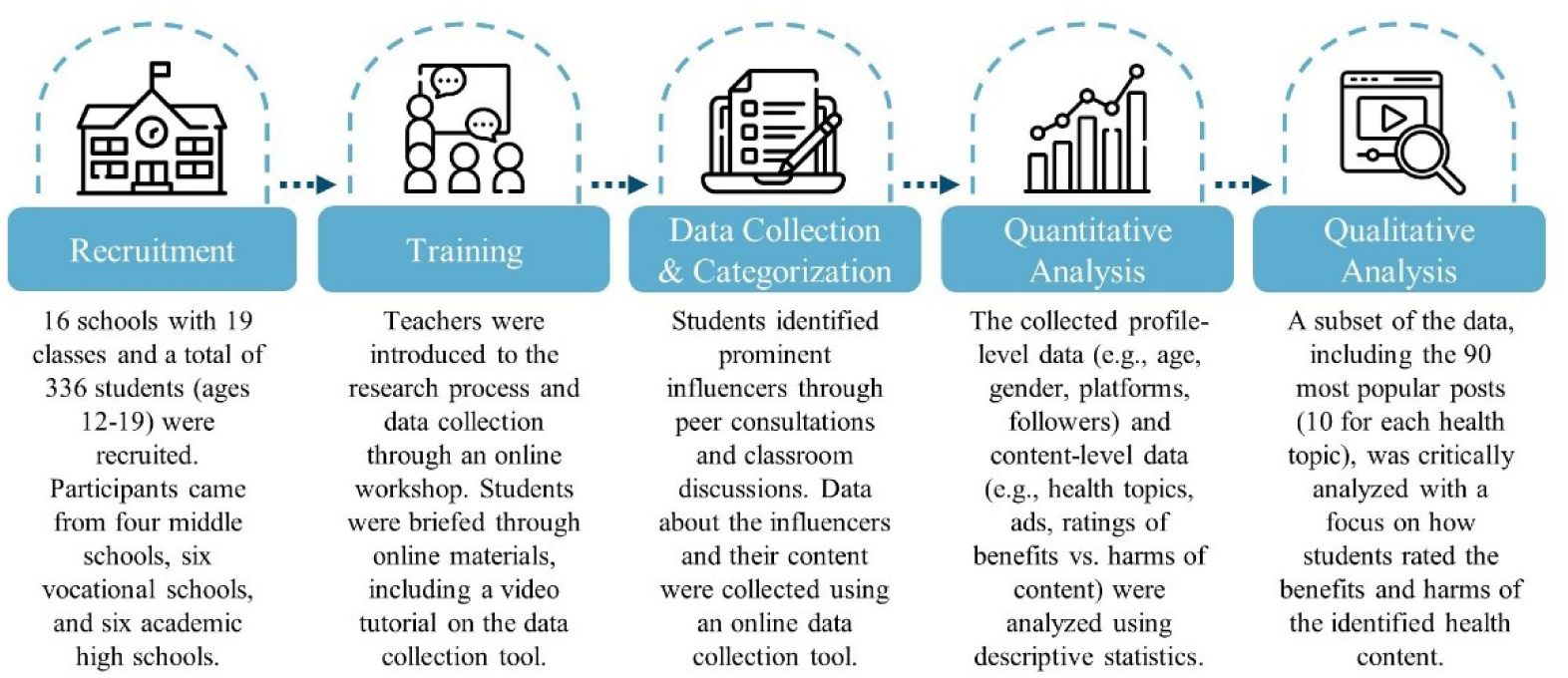
Youth-Driven Mixed Method Study

### 4.4 Content, Topics, and Disclosure of Advertisements

Advertisements were identified in 41.5% of all posts, primarily disclosed in the text (29.9%), logos (19.9%), and hashtags (15.1%), with verbal ad disclosures being the least common (12%) (Supplementary eTable 3).

The CYS documented 111 posts (5.2%) in the ‘other’ category, which include tagging companies or products in pictures or videos, providing affiliate links in captions, and influencers wearing or consuming branded products.

Most advertisements were identified in posts on supplements (84.1%), followed by addictive substances (55.6%), and food and nutrition (53.8%) (Figure 2). The least number of advertisements was identified in posts relating to beauty ideals (41.8%), sexual health (37.7%), and mental health (35.4%).

**Figure 2.**
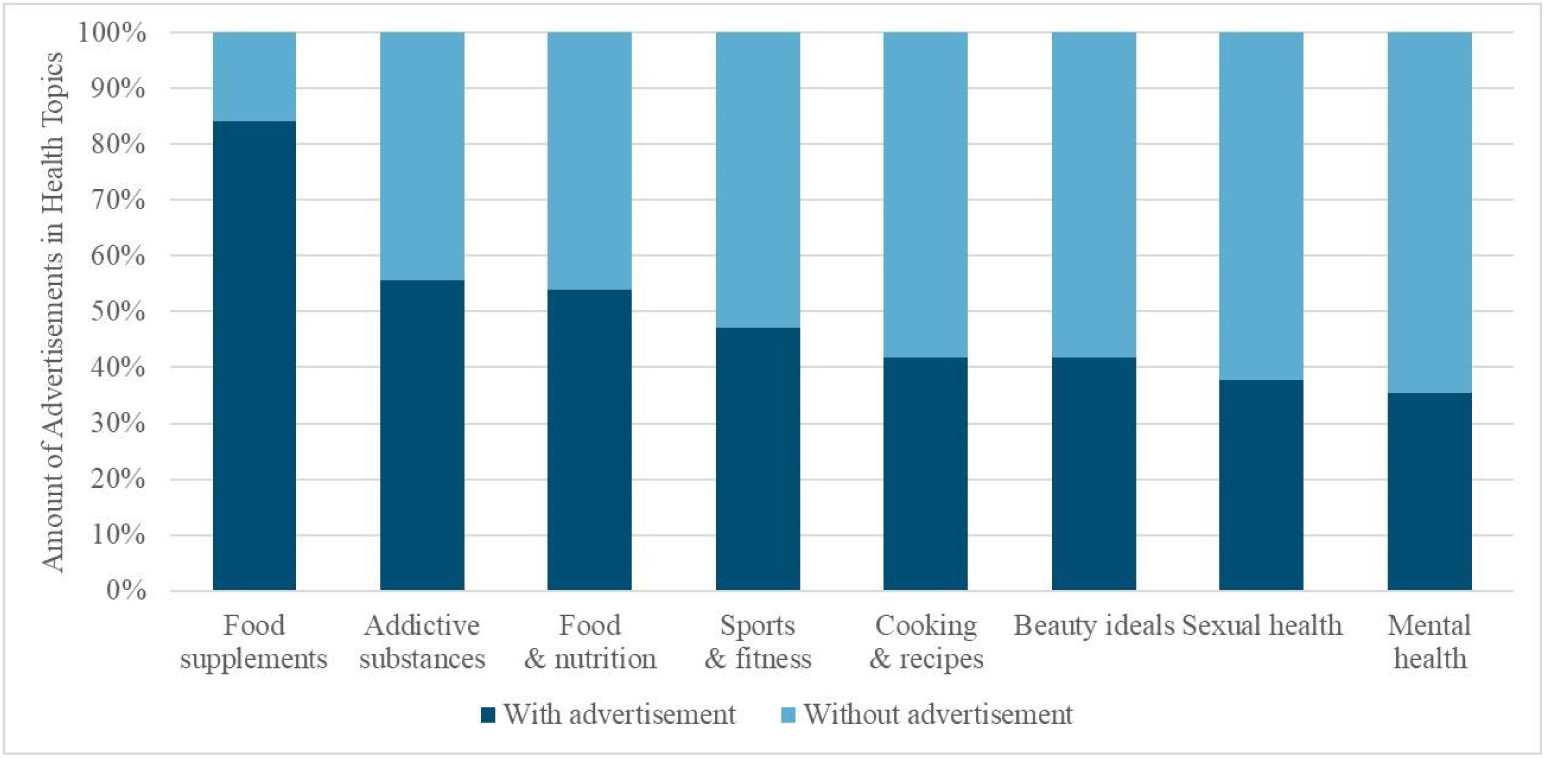
Percentage of posts containing advertising in the different health topics

The most frequently promoted products included foods (15.8%), beauty and lifestyle products (12.9%), supplements (11.2%), and sports equipment (10.8%). Only six posts (0.3%) advertised addictive substances, including cannabis in two and alcohol in four cases. It is also notable that only a minority of 22 posts (1.0%) contained undisclosed advertisements (Supplementary eTable 3).

Taking a closer look at adolescents’ health impact ratings reveals that the presence of advertisements had little impact on their ratings. Posts without advertisements were more often rated by the CYS as very beneficial (32.3%) and as somewhat harmful (5.2%), or very harmful (0.4%). In contrast, posts that included advertisements were more frequently rated as somewhat beneficial (38.9%) and as neither beneficial or harmful (34.9%) (Figure 3).

**Figure 3.**
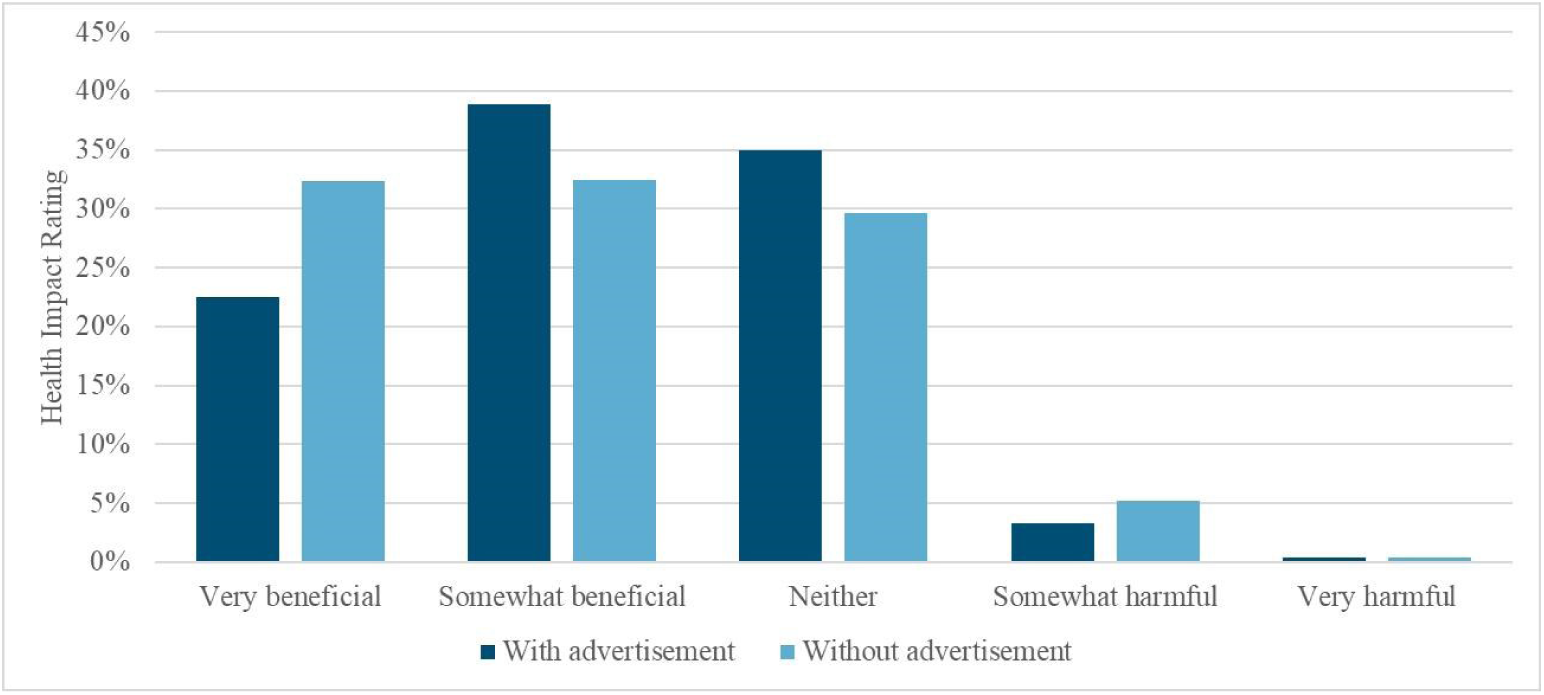
Health impact ratings from participating adolescents for posts with and without advertisements

## 5. Qualitative Results (Study 2)

Supplementary eTable 4 summarizes all information on the analyzed posts and advertisements, the ratings of adolescents and researchers, and the coding of potentially harmful content. When comparing the perceived health impact of influencer posts, the students perceived them as more beneficial for health than the research team. While the CYS rated 49 (54.4%) of the 90 posts as beneficial and 9 (10%) as harmful, the research team rated 18 posts (20%) as beneficial and 46 (51.1%) as harmful.

This discrepancy was particularly pronounced for the topics of food supplements, beauty ideals, cooking, and sports. The researchers categorized all posts related to food supplements as harmful, while adolescents rated only one post as harmful (which promoted an energy drink) and seven as beneficial. Six posts advertised online shops selling supplements (e.g., protein powder, vitamins, creatine), energy drinks, and ultra-processed foods (e.g., shakes, cookies, marmalades), and two posts advertised specific products (protein bar and nut butter). In contrast, the research team identified ultra-processed foods high in fat, sugar, and salt (Lawrence and Baker, 2019), as well as food supplements, may containing artificial sweeteners, undeclared pharmaceuticals, or steroids (Herriman et al., 2017; Or et al., 2019), as potentially harmful content. These promotions included misleading health claims, such as endorsing a questionable body ideal, centered on a lean, muscular, and sexually objectified physique (i.e., focus on abdominal, gluteal, or pectoral muscles).

In posts related to beauty standards, the research team rated nine as harmful and one as neutral, while the CYS rated four posts as beneficial and six as neutral. Three of the four posts rated as beneficial contained advertisements for ultra-processed foods, dietary supplements, clothing, and workout routines, all including sexually objectifying portrayals. Seven posts deemed harmful by the research team also included advertisements containing misleading health claims (i.e., consumption of promoted products leading to desired beauty standards).

Looking at posts related to cooking, the research team classified seven posts as harmful, while the CYS rated none as harmful. Influencers advertised potentially harmful products by using them while cooking (e.g., ultra-processed foods or supplements) or by presenting and consuming them during cooking (e.g., energy drinks, sweetened beverages, or alcohol). Problematic content included linking healthy nutrition to food supplements, associating athletic performance with energy drinks, and trivializing the consumption of alcohol.

While the CYS categorized none of the posts on sports, training, and fitness as harmful, the research team did so for six posts out of ten. These posts advertised food supplements and energy drinks, relating these products to unrealistic appearances and athletic performance. Physical activity was portrayed solely as a means of losing weight to reach a particular beauty standard (i.e., lean and muscular) without relating to other health benefits (e.g., cardiovascular health) (Warburton et al., 2006) or intrinsic motivations such as enjoyment, satisfaction, or sense of accomplishment (Teixeira et al., 2012).

Adolescents were most critical of content related to addictive substances and nutrition, rating four of ten posts as harmful. However, in the case of addictive substances, they considered four posts as neutral, although all of these posts either advertised or promoted the consumption of alcohol or drugs in a subtly way. For instance, in one post, the influencer seemingly advocates for the responsible use of drugs, but at the same time speaks about his positive experiences and promotes an online shop for drug derivatives.

Sexual and reproductive health received the most positive reception from both the CYS and the research team, with none of the documented posts categorized as harmful. None of the posts contained advertisements except for an intimate skincare product. Mental health content was also positively received, however, the researchers categorized two posts as harmful. These pictures and videos showed sexually objectifying portrayals and one video claiming to show “secret” symptoms of depression that can be deemed as common emotional reactions. Except for one post promoting a skincare product supporting the LGBTQIA+ community, none of the mental health posts included advertisements.

### 5.1 Examples of Youth Misconceptions

Table 1 presents one exemplary post for each health topic with high discrepancy in the health impact ratings (i.e., food supplements, beauty standards, cooking, and sports) to highlight potential misconceptions among adolescents. These posts were rated as beneficial by the adolescents but harmful by the researchers.

At first glance, all posts contain some beneficial content regarding physical activity and healthy nutrition. However, in all four cases, a healthy lifestyle is portrayed in conjunction with the consumption of advertised products. Furthermore, healthy nutrition and physical activity are promoted with a heavy focus on achieving an idealized and sexualized beauty image without addressing actual health benefits (e.g., cardiovascular health). Lastly, the posts include misleading health claims. For instance, a healthy diet needs to be accompanied by food supplements (Table 1).

## 6. Discussion

The sheer volume of advertisements (41.5% of all posts) identified in the influencers posts raises concerns and provides additional evidence that adolescents are prime targets for health-related marketing (Potvin et al., 2019; Montgomery and Chester, 2009). Furthermore, our findings revealed that influencers frequently promote unhealthy, untested supplements, or even addictive substances, such as alcohol or nicotine products. This is concerning, as young people often develop parasocial relationships with influencers, making them seem like relatable, authentic, friends or role models (Engel et al., 2024a). As a result, many young people look up to these influencers, which amplifies their impact on their behavior and perceptions (Giles, 2002; Yuan & Lou, 2020).

While the CYS in our study correctly identified a substantial amount of advertising, however, they still categorized most documented posts as beneficial for health. Rating content as beneficial in which influencers advertise unhealthy or untested products with misleading health claims or promote sexualized beauty ideals may indicate a lack of critical evaluation skills and persuasion knowledge (Friestad and Wright, 1994; Ku et al., 2019).

As shown in the exemplary posts (Table 1), influencer posts appear to contain beneficial health messages at first glance. However, a closer look reveals concerning content, including sexualized beauty ideals and misleading health claims, that have been overlooked by the adolescents. This suggests that adolescents may have a good grasp of basic healthy habits, such as staying active and eating well, while they lack the media literacy and critical reflection skills required to navigate the complex and commercialized digital environment (Buckingham, 2008; Naderer, 2021).

Helping adolescents develop critical thinking skills and persuasion knowledge is essential (Wang and Togher, 2024). These skills enable them to recognize subtle persuasive tactics and evaluate content more critically. A key part of the solution lies in school education. In many countries, including Austria, young people lack sufficient digital health literacy to understand and assess the unmoderated and often misleading health messages they encounter online (Heiss et al., 2025). Therefore, it is important to revise school curricula to integrate health education more thoroughly, either as a standalone subject or a cross-curricular theme.

It is also important to note that the health-related content documented by CYS is significantly shaped by prominent women, typically aged between 19 and 39, who mainly use Instagram. The popularity of young female influencers among the participating CYS raises concerns about the potentially unequal influence exerted on adolescent girls due to gender-specific effects. For instance, research showed that women, compared to men, were more likely to perceive themselves as similar to influencers, resulting in stronger feelings of para-social interaction and, in turn, positively impacting brand attitude towards advertised products (Hudders and De Jans, 2022). Moreover, considering the sexualized and objectified content of numerous posts, it is expected that girls will be more adversely affected by the negative outcomes of self-objectification (Karsay, 2020).

Lastly, only a few posts treated specific illnesses (e.g., cancer, cellulite, endometriosis, lipedema). Although these illnesses may not be as marketable as other health topics, influencers still provide information on various diseases and may thus function as important information hubs on these more serious health issues. However, celebrity endorsements on social media, often lacking scientific grounding and driven by financial interests, can spread misleading health information, posing particular risks for vulnerable groups such as cancer patients (Gram et al., 2025). This underscores the need to investigate more closely how accurately and in which contexts influencers discuss diseases.

### 6.1 Limitations

Even though this is the first study, to our knowledge, that seeks to open the black box of influencers’ health content by employing a participatory citizen science approach, the study also has some limitations. First, the selection of influencers and posts was based on the choices of participating CYS, which introduces a bias toward popular celebrity influencers and may not fully represent the diversity of health-related content encountered in personalized social media environments. Second, the subjective nature of the evaluations provided by the CYS poses another limitation. Despite careful training, the adolescents’ assessments of health impact and advertisement density may reflect personal biases or limited understanding of the content’s implications. This subjectivity underscores the importance of complementing citizen science approaches with expert evaluations, as demonstrated in Study 2, to ensure a more balanced and accurate analysis. Finally, the study’s reliance on self-reported data and observations from adolescents limits its ability to capture the full complexity of algorithmic curation and click behavior within social media platforms. Future studies should aim to integrate data from platform analytics to better understand how adolescents interact with influencer content and how algorithms shape their exposure to health-related posts. Addressing these limitations will strengthen the coherence and impact of future research in this field.

## 7. Conclusion

The study revealed that influencers’ health content is dominated by advertisements, often promoting ultra-processed food, untested nutrition supplements, idealized beauty ideals, or undesirable health behavior, including the consumption of addictive substances. The findings of this study underscore the critical need for educational interventions aimed at enhancing adolescents’ critical thinking and media literacy. Study 2 revealed significant misconceptions among adolescents regarding the health content shared by influencers, particularly in their inability to discern between genuine health information and content influenced by commercial motives. These misconceptions highlight gaps in adolescents’ persuasion knowledge and their ability to critically evaluate the credibility and intent of health-related social media posts. To address these gaps, it is essential to integrate digital health literacy into school curricula, either as a standalone subject or as part of a cross-curricular theme. Such education should focus on equipping young people with the skills to recognize persuasive tactics, evaluate the credibility of health information, and identify misleading claims. By fostering these competencies, adolescents will be better prepared to navigate the complex and commercialized digital health environment effectively. Furthermore, stricter regulations on harmful and misleading content promoted by influencers are necessary to safeguard youth from exposure to potentially detrimental health messages.

## Acknowledgements

We thank the students from the participating middle and high schools for their contributions to data collection and categorization in this study. We also thank Steven Woloshin, MD, for his valuable feedback

## Ethics Statement

Ethical approval was obtained from the IRB of the MCI | The Entrepreneurial School® (reference number: 20230312)

## Data Availability Statement

The datasets generated and analyzed during the current study are available from the corresponding author upon request.

## Declaration of generative AI

During the preparation of this work, the authors used ChatGPT and DeepL to check grammar and improve readability. The authors reviewed and edited the content as needed and take full responsibility.

