## Supplemental Material for "Uncovering Social Media Influencers’ Health Posts: A Youth-Driven Mixed Method Study"

### Uncovering Social Media Influencers' Health Content: A Citizen Youth Science Study

#### Supplementary Material

**eTable 1.** Data collection tool…………………………………………………………………………………...2

**eTable 2.** Profile-level data…………………………………………………………………………………...9

**eTable 3.** Post-level data……………………………………………………………….................................10

**eTable 4.** Qualitative content analysis of influencer posts………………………………………………………...13

**Table A1.** Data collection tool

| Profile Data | | |
|---|---|---|
| Plattforms, Name(s) and Followers of the Influencers<br>Please click the relevant box and fill in the Names and Followers | | |
| YouTube | Instagram | TikTok |
| ☐ | ☐ | ☐ |
| Name on YouTube: | Name on Instagram: | Name on TikTok: |
| Followers on YouTube: | Followers on Instagram: | Followers on TikTok: |

| About which health topics does the influencer post?<br>Please click the relevant box | | | | | |
|---|---|---|---|---|---|
| Food & Nutrition | Sport & Fitness | Mental Health | Sexuality | Addictive Substances (e.g. Alcohol, Tobacco) | Other topics |
| ☐ | ☐ | ☐ | ☐ | ☐ | ☐ |
| Other topics: Please type in any other topics here | | | | | |

| Which language does the influencer mainly speak in the posts?<br>Please click the relevant box |
|---|

| German | English | Other language |
| --- | --- | --- |
| ☐ | ☐ | ☐ |

Other language: Please type in the other language here

Is the influencer of any gender?
Please click the relevant box

| Male | Female | Trans, non-binary, diverse | Don't know |
| --- | --- | --- | --- |
| ☐ | ☐ | ☐ | ☐ |

Which country does the influencer come from?
Please click the relevant box

| Austria, Germany or Switzerlandz | Other countries in Europe | USA, Canada or Australia | Other |
| --- | --- | --- | --- |
| ☐ | ☐ | ☐ | ☐ |

How old is the influencer?

| | |
|---|---|
| Please type the age as a number here | Don't know |
| | FALSE |

**Post Data**

Is there a title for the post?

If available, please enter title here

When was the post published?

Please type in the publication date of the post here

Please provide a link to the post!

Please insert the link to the post here

On which platform was the post uploaded?
Please click the relevant box

| | | |
|---|---|---|
| YouTube | Instagram | TikTok |
| ☐ | ☐ | ☐ |

| | | | |
|---|---|---|---|
| What is the format of the post? Is there any text, video, image or a audio?<br>Please click the relevant box | | | |
| 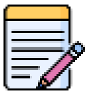 | 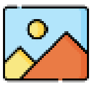 | 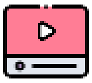 | 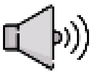 |
| Text | Image | Video | Audio/Sound |
| ☐ | ☐ | ☐ | ☐ |

| | | | |
|---|---|---|---|
| What health topics are mentioned in the article/post?<br>Please click the relevant box | | | |
| 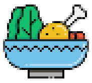 | 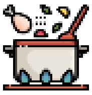 | 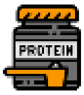<br> | 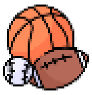 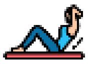 |
| Food & Nutrition | Cooking & Recipes | Food supplements | Sport, Training & Fitness |
| ☐ | ☐ | ☐ | ☐ |
| 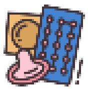 | 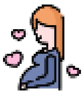 | 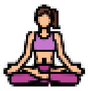 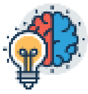 | 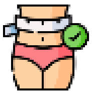 |
| Sexual Health (e.g. Sexually transmitted diseases) | Reproductive Health (e.g. Pregnancy) | Mental Health | Beauty Ideals (e.g. thin, muscular) |
| ☐ | ☐ | ☐ | ☐ |
| 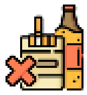 | 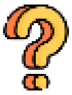 | Are there any other health topics that the influencer adresses?<br>Please type in here | |
| Addictive Substances (e.g. Vaping, Alcohol, Cannabis) | Other health topics | | |

| | |
|---|---|
| ☐ | ☐ |

| |
|---|
| Are any specific diseases mentioned in the post?<br>Please click the relevant box |

| | |
|---|---|
| Yes | No |
| ☐ | ☐ |

| |
|---|
| The following diseases occur in the post: Please insert the disease(s) here |

| |
|---|
| Is the content of the post beneficial or harmful for a healthy life?<br>Please click the relevant box |

| | | | | | |
|---|---|---|---|---|---|
| Very beneficial | Somewhat beneficial | Neither | Somewhat harmful | Very harmful | Don't know |
| ☐ | ☐ | ☐ | ☐ | ☐ | ☐ |

| |
|---|
| Is there any advertising (e.g. paid content, collaborations or sponsorship) in this post?<br>Please click the relevant box |

| | | |
|---|---|---|
| Yes | No | Don't know |
| ☐ | ☐ | ☐ |

In what way is advertising or paid content indicated in this post?
Please click the relevant box

| No indicator | Hashtag | In the text | In the picture (e.g. Coca Cola logo) | Mentioned by the influencer | Other indicators |
|---|---|---|---|---|---|
| ☐ | ☐ | ☐ | ☐ | ☐ | ☐ |

If there are other indicators: Please type in any other clue/indicator here

Which products are advertised in the post?
If no advertising, skip this question

| Addictive substance | Sports equipment | Food & Nutrition | Beauty & Lifestyle |
|---|---|---|---|
| ☐ | ☐ | ☐ | ☐ |
| Technical devices | Apps (e.g. Spotify, Freeletics) | Workshop or workout program | Other products |
| ☐ | ☐ | ☐ | ☐ |

Other advertised products: Please type in any other products here

How popular is this post?

Please insert the number as a numerical value (e.g. 150, 300) or leave the field blank if not available

| Likes | Comments | Shared | Views |
|---|---|---|---|
| Please insert number here | Please insert number here | Please insert number here | Please insert number here |

**Table A2.** Profile-level data

| | |
|---|---|
| **Documented Profiles** | 255 (100%) |
| **Platform** (multiple response) | |
| YouTube profiles | 186 (72.9%) |
| Instagram profiles | 243 (95.3%) |
| TikTok profiles | 209 (81.9%) |
| **Influencer type** | |
| Micro (1.000 - 20.000) | 14 (5.5%) |
| Meso (20.000 - 100.000) | 26 (10.2%) |
| Macro (100.000 - 300.000) | 42 (16.5%) |
| Celebrity (300.000+) | 172 (67.5%) |
| **Platforms used** | |
| 1 Platform | 29 (11.4%) |
| 2 Platforms | 69 (27%) |
| 3 Platforms | 157 (61.6%) |
| **Language** (multiple response) | |
| English | 109 (42.8%) |
| German | 164 (64.3%) |
| Other | 15 (5.9%) |
| **Gender** | |
| Male | 103 (40.4%) |
| Female | 143 (56.1%) |
| Trans, non-binary, diverse | 0 (0%) |
| Not documented | 9 (3.5%) |
| **Country** | |
| Austria, Germany, Switzerland | 157 (61.5%) |
| Other countries in Europe | 25 (9.8%) |
| USA, Canada, Australia | 56 (22%) |
| Other | 12 (4.7%) |
| Not documented | 5 (2%) |
| **Age** | |

| | |
|---|---|
| 14-18 | 6 (2.4%) |
| 19-29 | 97 (38%) |
| 30-39 | 58 (22.7%) |
| 40-49 | 5 (2%) |
| 50-59 | 5 (2%) |
| 60+ | 0 (0%) |
| Not documented | 84 (32.9%) |
| **Profile focus** (multiple response) | |
| Food & nutrition | 158 (62%) |
| Sport & fitness | 154 (60.4%) |
| Mental health | 50 (19.6%) |
| Sexual health | 32 (12.6%) |
| Substance use | 10 (3.9%) |
| Other | 60 (23.5%) |

**Table A3.** Post-level data

| **Documented Posts** | 2147 (100%) |
|---|---|
| **Platform** | |
| YouTube | 402 (18.7%) |
| Instagram | 1213 (56.5%) |
| TikTok | 517 (24.1%) |
| Not documented | 15 (0.7%) |
| **Format** (multiple response) | |
| Text | 1557 (72.5%) |
| Picture | 697 (32.6%) |
| Audio | 1331 (62%) |
| Video | 1585 (73.8%) |
| Not documented | 15 (0.7%) |
| **Health topic** (multiple response) | |
| Food & nutrition | 593 (27.6%) |
| Cooking & recipes | 410 (19.1%) |

| | |
|---|---|
| Food supplements | 160 (7.5%) |
| Sports, training & fitness | 1084 (50.5%) |
| Sexual health | 29 (1.4%) |
| Reproductive health | 41 (1.9%) |
| Mental health | 258 (12%) |
| Beauty ideals | 635 (29.6%) |
| Addictive substances | 20 (0.9%) |
| Others | 144 (6.7%) |
| Not documented | 19 (0.9%) |
| **Illness mentioned** | |
| Yes | 104 (5.5%) |
| No | 2001 (92.5%) |
| Not documented | 42 (2%) |
| **Health impact** | |
| Very beneficial | 578 (26.9%) |
| Somewhat beneficial | 719 (33.5%) |
| Neither | 651 (30.3%) |
| Somewhat harmful | 87 (4.1%) |
| Very harmful | 8 (0.4%) |
| Not documented | 104 (4.8%) |
| **Ad** | |
| Yes | 891 (41.5%) |
| No | 1114 (51.9%) |
| Not documented | 142 (6.6%) |
| **Ad type** (multiple response) | |
| No disclosure | 482 (22.5%) |
| Hashtag | 325 (15.1%) |
| Text (post or video) | 621 (28.9%) |
| Picture or logo | 427 (19.9%) |
| Verbal (audio or video) | 258 (12%) |
| Others | 111 (5.2%) |

| | |
|---|---|
| Not documented | 14 (0.7%) |
| **Product type** (multiple response) | |
| Addictive substances | 6 (0.3%) |
| Sports equipment | 232 (10.8%) |
| Food & nutrition | 339 (15.8%) |
| Food supplements | 240 (11.2%) |
| Technical gadgets | 52 (2.4%) |
| Apps | 88 (4.1%) |
| Workshop & training | 140 (6.5%) |
| Beauty & lifestyle | 276 (12.9%) |
| Not documented | 14 (0.7%) |
| **Engagement** (like + comment) | |
| 1. Quartil | 3310,5 |
| 2. Quartil | 13487 |
| 3. Quartil | 55719 |
| 4. Quartil (Maximum) | 13291689 |

**Table A4.** Qualitative content analysis of influencer posts

| Health Topic | Influencer Profile | **Post Engagement** (likes & comments) | Post Description | Classification of Citizen Scientists | Classification of Research Team | Advertising Content | Problematic Content |
|---|---|---|---|---|---|---|---|
| Addictive Substances | Instagram: miakhalifa<br><br>Follower: 37.300.000 | 1.875.102 | Several sexualized lifestyle pictures of the influencer.<br>The last picture depicts cannabis use and cannabis products from the influencers product line (Khalifa Kush). | Neutral | Harmful | Clothing labels (1xblue, tommyjeans, stellamccartney)<br><br>Restaurants (hiyakawamiami)<br><br>Books (The Idiot, The Bitch)<br><br>Cannabis product (Khalifa Kush) | Consumption and presentation of cannabis products. Potentially harmful beauty ideals (i.e. sexualized, thin, muscular) |
| Addictive Substances | Instagram: rezo<br><br>Follower: 1.200.000 | 138.825 | Picture depicting an intoxicated influencer sitting on the ground (probably in front of a public toilet). | Harmful | Harmful | Red Bull cup | Humorous capture accompanying the post: “partying again after 2 years". Connecting partying and fun with the consumption of addictive substances. |
| Addictive Substances | TikTok: thegeneticone<br><br>Follower: 1.100.000 | 84.892 | Video of topless fitness influencer flexing his muscles with captions. I.e.:“you should be on stage”.<br><br>This is followed by a short video montage showing the influencer drinking alcohol on several occasions (pubs, festivals etc.) | Neutral | Harmful | Gym (Iron Gym)<br><br>Alcohol brands (Gösser, Eristoff)<br><br>Protein Powder | Humorous montage showing fun activities (festivals, games, mingling with friends, clubbing) in connection with the consumption of alcohol. |
| Addictive Substances | YouTube: Michael Smolik<br><br>Follower: 811.000 | 57.025 | Video of a professional martial artist recounting a story about self-defense when he was attacked by a drunk while walking home with friends. Although, he repeatedly<br>However, he repeatedly emphasizes to avoid violent conflicts. | Neutral | Neutral | Clothing of own product line (Team Smolik)<br><br>Trainings, Seminars, Boot Camps | The influencer points out that they were attacked by "foreign citizens" and repeatedly describes them as “asocial”. |
| Addictive Substances | YouTube: OPEN MIND<br><br>Follower: 538.000 | 45.109 | The video is an experience report by the influencer about his first cocaine use and its side effects. | Beneficial | Harmful | Clothing of own product line<br><br>Promotion of influencers online shop (gadgets for | Although the influencer repeatedly advises against drug use, his experiences are presented in a humorous manner, and on several |

| | | | | | | | |
|---|---|---|---|---|---|---|---|
| | | | He repeatedly advises against drug use and describes his actions in the video for educational purposes. | | | consumption, drug derivatives, food supplements) | occasions, tips for purchasing and consumption are given. |
| Addictive Substances | YouTube: Brittany Broski<br><br>Follower: 538.000 | 44.578 | The influencer video shares recipes of prohibition cocktails from the advertised book and tests them. | Harmful | Harmful | Alcohol brands (Amore, Cointreau, Kahlua, Tuca, Tito's)<br><br>Sweetend Beverages (Pepsi, Goslings, Simply Orange; Rose's)<br><br>Food (Dole, tipsy)<br><br>Book (Drink Like a Mobster) | Promotion of alcohol consumption with humorous and positive descriptions (i.e. delicious, affordable) |
| Addictive Substances | TikTok: thegeneticone<br><br>Follower: 1.100.000 | 32.500 | This influencer video provides tips on balancing drinking and partying with maintaining a training/gym schedule. | Neutral | Harmful | Alcohol brands (Havana Club, Berliner Luft, Smirnoff,AL GE, Corona )<br><br>Food Supplements (electrolytes, minerals) | The video suggests that with the influencer's tips the negative consequences of alcohol can be counteracted and the consumption is trivialized. |
| Addictive Substances | Instagram: thehealchef<br><br>Follower: 610.000 | 21.980 | The influencer's post contains several lifestyle pictures accompanied by an extensive caption explaining why he chose to go sober for one year. He criticizes his cultural upbringing, where excessive alcohol consumption is accepted and encouraged. Furthermore, they recount all the positive changes in their life that occurred due to sobriety. | Beneficial | Beneficial | No ads | |
| Addictive Substances | YouTube: OPEN MIND<br><br>Follower: 538.000 | 21.296 | The video is an experience report by the influencer about his first consumption of the then legal THC derivatives HHC. | Harmful | Harmful | Promotion of influencers online shop (gadgets for consumption, drug derivatives, food supplements) | Although the influencer repeatedly advises against drug use, yet his anecdotes are humorously framed, offering advice on acquiring and using drugs. Additionally, he promotes HHC as a THC alternative that won't trigger positive results on drug tests. |
| Addictive Substances | YouTube: Manuel Haas<br><br>Follower: 93.700 | 10.248 | The video is an experience report by the influencer about his first consumption of cannabis. | Harmful | Harmful | Food supplements (rockanutrition) | The consumption is presented humorously and positively framed. Furthermore, no risks, age restrictions or advice against drug use are given. |

| | | | | | | | |
|---|---|---|---|---|---|---|---|
| Beauty Ideal | YouTube:<br>Cloe Ting<br><br>Follower:<br>24.500.000 | 4.049.395 | The video shows the influencer doing a whole body workout. Furthermore, the influencer promises that following this workout for two weeks leads to considerable weight loss. | Beneficial | Harmful | Clothing (own product line Cloeting)<br><br>Workouts (own series) | The video suggests that one should work out to lose weight and not other health benefits. Promotion of own workouts and product line for best results. Promotion of questionable beauty ideal (i.e. thin, muscular, “shredded”) |
| Beauty Ideal | Instagram:<br>kyliejenner<br><br>Follower:<br>389.000.000 | 3.694.523 | Several pictures showing the influencer posing with her new makeup products (mascara, liquid eyeshadow) | Neutral | Harmful | Makeup (kylie cosmetics) | Promotion of questionable beauty ideal (i.e. thin, muscular, wearing makeup) |
| Beauty Ideal | Instagram:<br>kyliejenner<br><br>Follower:<br>389.000.000 | 3.250.944 | The picture shows the influencer posing with a water bottle in sportswear. | Beneficial | Harmful | Water (Glow) | Promotion of a questionable beauty ideal (i.e. thin, muscular, sexualized). Promotion of water with artificial products. |
| Beauty Ideal | Instagram: miakhalifa<br><br>Follower: 37.300.000 | 2.631.149 | Several pictures of the influencer posing in a costume with high skin exposure. | Neutral | Harmful | Clothing (Nippies, hallofframes, Oakley) | Promotion of sexualized and questionable beauty ideal (i.e. thin, muscular, skin exposure) |
| Beauty Ideal | Instagram:<br>cbum<br><br>Follower:<br>17.000.000 | 1.798.076 | Several pictures of the influencer posing and flexing his muscles in a gym, partly while working out. | Beneficial | Harmful | Clothing (youngla)<br><br>Food supplements (getrawnutrition) | Promotion of a questionable beauty ideal (i.e. muscular) in conjunction with food supplements. |
| Beauty Ideal | Instagram: miakhalifa<br><br>Follower: 37.300.000 | 1.266.477 | The video and two pictures show the influencer posing | Neutral | Harmful | Clothing (bodiedbyuchis) | Promotion of questionable and sexualized beauty ideal (i.e. thin, muscular, skin exposure) |
| Beauty Ideal | Instagram:<br>cbum<br><br>Follower:<br>17.000.000 | 861.910 | The picture shows the influencer posing and flexing with his coach. | Beneficial | Neutral | No ads | - |
| Beauty Ideal | Instagram:<br>davidlaid<br><br>Follower:<br>3.300.000 | 817.819 | Several pictures and selfies of the influencer posing in a gym and flexing his muscles while promoting a clothing brand. | Neutral | Harmful | Clothing (gymshark) | Promotion of a questionable beauty ideal (i.e. thin, muscular). |

| | | | | | | | |
|---|---|---|---|---|---|---|---|
| Beauty Ideal | Instagram: davidlaid<br><br>Follower: 3.300.000 | 541.365 | Picture depicting the influencer in a bathtub with exposed upper body. | Neutral | Harmful | No ads | Promotion of a questionable and sexualized beauty ideal (i.e. muscular, exposed) |
| Beauty Ideal | Instagram: davidlaid<br><br>Follower: 3.300.000 | 525.712 | The picture shows the influencer posing and flexing his muscles while his upper body is exposed. | Neutral | Harmful | No ads | Promotion of a questionable and sexualized beauty ideal (i.e. muscular, exposed) |
| Cooking | Instagram: fitwafflekitchen<br><br>Follower: 2.100.000 | 2.329.000 | The video shows how to cook and fry french toast sticks with nutella stuffing. | Beneficial | Harmful | Nutella | Promotion of highly processed foods high in saturated fats and sugar. |
| Cooking | Instagram: simpyni_<br><br>Follower: 1.200.000 | 2.000.000 | The video shows in a highly stylized way the preparation of pancakes and a cofé with different liquors. | Neutral | Harmful | No ads | Pancakes high in fat and sugar as well as the promotion of alcoholic beverages. |
| Cooking | YouTube: KWOOK<br><br>Follower: 3.800.000 | 594.920 | YouTube video where the influencer informs about Japanese Christmas traditions. The video suggests that KFC (Kentucky Fried Chicken) has had a central role since the 70s. According to the video, eating a bucket of KFC fried chicken is the most popular Christmas tradition among the Japanese. Furthermore, the influencer is shown eating fried chicken and fries. | Neutral | Harmful | Fast Food Chain (Kentucky Fried Chicken) | Promotion of highly processed fast food high in saturated fats and sugar. |
| Cooking | Instagram: esmuellert<br><br>Follower: 12.800.000 | 331.827 | Pictures depicting the influencer preparing spinach, accompanied by a caption with information on the benefits and contents of spinach. | Beneficial | Beneficial | No ads | - |
| Cooking | YouTube: KWOOK<br><br>Follower: 3.800.000 | 283.702 | The video shows the influencer explaining a cheap recipe to cook pasta. | Neutral | Neutral | Cookbook from the influencer | - |

| | | | | | | | |
|---|---|---|---|---|---|---|---|
| Cooking | YouTube: KWOOK<br><br>Follower: 3.800.000 | 264.274 | YouTube video showing the influencer eating hot dogs from a fast food chain (Pink's Hot Dogs) and rating them. | Neutral | Harmful | Fast Food Chain (Pink's Hot Dogs) | Promotion of highly processed fast food high in saturated fats and sugar. |
| Cooking | Instagram: leanadeep<br><br>Follower: 4.700.000 | 213.179 | The influencer is portrayed in a video demonstrating the preparation of breakfast, lunch, and dinner, with captions detailing the recipes. However, the meal preparation is often interrupted by the consumption of protein powder drinks and training sequences. | Beneficial | Harmful | Food Supplements from influencers own product line (bloom greens, bloom tiger's blood) | Promotion of food supplements suggests that healthy nutrition while working out must be accompanied by the consumption of these supplements. |
| Cooking | Instagram: katjakrasavice | 192.139 | Several pictures showing the influencer sitting in a warehouse on pallets of cans of her new line of soft drinks. | Neutral | Harmful | Soft drinks (sugarmami) | Promotion of sweetened beverages high in sugar. |
| Cooking | YouTube: Doctor Mike<br><br>Follower: 11.900.000 | 191.200 | In the video the influencer tests the keto diet for 30 days. He shows his meal preparations, workouts and day to day life. He explains the idea behind this diet and informs about the pros and cons. | Neutral | Neutral | No ads | - |
| Cooking | TikTok: leanadeep<br><br>Follower: 12.400.000 | 183.079 | The video shows the influencer preparing different meals for several days. In the caption the recipes are provided. | Neutral | Harmful | Food Supplements (bloom greens) | Promotion of food supplements suggesting that preparing healthy meals must be accompanied by food supplements. |
| Food Supplements | Instagram: daisykeech<br><br>Follower: 5.800.000 | 393.222 | Picture of an influencer posing in sportswear with an energy drink. | Harmful | Harmful | Energy drink (Celsius) | Depiction of a questionable body type (i.e. thin, muscular, sexualized) and promotion of energy drink. |
| Food Supplements | Instagram: aliciasmd<br><br>Follower: 3.700.000 | 352.014 | Picture of an influencer posing in sportswear and promoting an online shop for food supplements. | Beneficial | Harmful | Food supplements (foodspring) | Depiction of a questionable beauty ideal (i.e. thin, muscular, sexualized) and promoting highly processed foods and supplements. |
| Food Supplements | Instagram: alex_eubank31<br><br>Follower: 2.300.000 | 342.994 | Several pictures of an influencer showing his body transformation while promoting several products. | Beneficial | Harmful | Clothing (Elysium, Ragwear)<br><br>Food supplements and energy drink (Alpha Lion, Celsius) | Depiction of questionable beauty ideals (i.e. thin, muscular) with the promotion of processed foods and questionable food supplements. |

| | | | | | | | |
|---|---|---|---|---|---|---|---|
| | | | | | | Ready-to-eat meals (FlexPro meals) | |
| Food Supplements | Instagram: pamela_rf<br><br>Follower: 9.100.000 | 243.402 | Picture of an influencer posing with a muesli bar while holding her dog. | Neutral | Harmful | Muesli bars (HEJnatural) | Depiction of questionable beauty ideals (i.e. thin, muscular) and promotion of sugary muesli bars as healthy snacks. |
| Food Supplements | Instagram: aliciasmd<br><br>Follower: 3.700.000 | 210.309 | Picture of an influencer posing in sportswear and promoting an online shop for food supplements. | Beneficial | Harmful | Food supplements (foodspring) | Depiction of a questionable beauty ideal (i.e. thin, muscular, sexualized) and promoting highly processed foods and supplements. |
| Food Supplements | Instagram: daisykeech<br><br>Follower: 5.800.000 | 202.940 | Picture of an influencer posing in sportswear with an energy drink. | Neutral | Harmful | Energy drink (Celsius) | Depiction of a questionable body type (i.e. thin, muscular, sexualized) and promotion of energy drink. |
| Food Supplements | Instagram: rezo<br><br>Follower: 1.200.000 | 196.981 | Pictures of an influencer showing his body transformation exposing his upper body. | Beneficial | Harmful | Food supplements and foods (morenutrition) | Promotion of highly processed foods and food supplements (high in sugar and preservatives). |
| Food Supplements | Instagram: pamela_rf<br><br>Follower: 9.100.000 | 148.550 | Several pictures of the influencer posing with with own food product (nut butter). | Beneficial | Harmful | Nut butter (naturally PAM) | Promoting processed food which is high in sugar and fats. |
| Food Supplements | TikTok: michikleis<br><br>Follower: 179.900 | 145.636 | Video of an influencer who reports on the advantages and disadvantages of magnesium on the basis of two studies. | Beneficial | Harmful | No ads | General promotion of supplement without context (i.e. age, gender, health condition). |
| Food Supplements | Instagram: pamela_rf<br><br>Follower: 9.100.000 | 131.839 | Several pictures and videos of the influencer with her partner posing and testing protein bars of their own product line. | Beneficial | Harmful | Protein bar (naturally PAM) | Promoting processed food which is high in sugar and fats implying it is a healthy alternative to chocolate bars. |
| Mental Health | Instagram: growingannanas<br><br>Follower: 832.000 | 1.798.462 | The video depicts the influencer humorously portraying various stages of emotions, motivations, and energy levels corresponding to the four stages of ovulation. | Beneficial | Beneficial | No ads | - |
| Mental Health | Instagram: coupleontour | 1.123.400 | The post contains two pictures of an influential couple holding a baby. The caption to the post | Beneficial | Neutral | No ads | - |

| | | | | | | | |
|---|---|---|---|---|---|---|---|
| | Follower:<br>2.500.000 | | tells the story of one mother having narrowly survived a stroke shortly before birth. | | | | |
| Mental Health | Instagram:<br>coupleontour<br><br>Follower:<br>2.500.000 | 994.015 | The post contains two pictures showing an influencer couple holding a skin care product from nivea and one picture showing only the nivea product with rainbow colors. With the post they suggest that nivea supports the LGBTQ+ community. | Beneficial | Neutral | Skin care (nivea) | While the influencers demonstrate their support for the LGBTQ+ community, it raises significant questions to link this support directly to a brand as part of their marketing strategy. |
| Mental Health | TikTok:<br>leanadeep<br><br>Follower:<br>12.400.000 | 435.481 | The video shows the influencer exercising when she unexpectedly gets a fist bump and a compliment from a stranger in the gym for her perseverance. The video includes the quote “little acts of kindness can change someone’s whole day”. | Beneficial | Neutral | No ads | - |
| Mental Health | YouTube:<br>thewizardliz<br><br>Follower:<br>4.160.000 | 433.711 | The video shows the influencer giving advice on how to change one's life for the “better”. | Neutral | Neutral | Apps (InShot, Pinterest) | - |
| Mental Health | YouTube:<br>thewizardliz<br><br>Follower:<br>4.160.000 | 408.345 | The video shows the influencer giving advice on relationships. | Beneficial | Neutral | No ads | - |
| Mental Health | Instagram:<br>growingannanas<br><br>Follower:<br>832.000 | 381.473 | The video depicts the influencer receiving negative comments about her makeup and how she deals with them. She promotes in her caption self-esteem among young women and counteracts beauty ideals. | Beneficial | Beneficial | No ads | - |
| Mental Health | Instagram:<br>pamela_rf<br><br>Follower:<br>9.100.000 | 275.672 | The video and several pictures show the influencer on diamond beach, where she was conceived by her parents. | Neutral | Harmful | No ads | Sexualized pictures and videos promoting a questionable body type (i.e. thin, muscular, sexualized). |
| Mental Health | TikTok:<br>vivis.psychologie<br><br>Follower: | 246.166 | The video shows the influencer talking about five secret signs of depression backed with music. | Beneficial | Harmful | No ads | Although the video is informative, the five can also be normal emotional states, especially in teenage years. |

| | | | | | | | |
|---|---|---|---|---|---|---|---|
| | 179.100 | | | | | | Questionable information like in this video can invite self-diagnosis. |
| Mental Health | TikTok: growingannanas<br><br>Follower: 1.000.000 | 242.939 | The video depicts the influencer giving advice on habits (i.e. workouts, cold showers, posture) which makes one feel good and powerful. | Beneficial | Neutral | No ads | The video suggests that these tips help universally for a strengthened self-confidence. |
| Nutrition | Instagram: kyliejenner<br><br>Follower: 389.000.000 | 2.815.680 | The picture shows four cups of take away coffee. | Harmful | Neutral | Makeup (Kylie Cosmetics)<br><br>Skincare (Kylie Skin)<br><br>Café (Alfreds | - |
| Nutrition | TikTok christian.wolf<br><br>Follower: 580.100 | 337.283 | The influencer debunks misinformation about coconut sugar promoted by another influencer (pamela_rf). | Beneficial | Beneficial | No ads | |
| Nutrition | YouTube: Sascha Huber<br><br>Follower: 1.650.000 | 333.794 | The Influncer describes in his video how you can create a "perfect" nutrition plan for your dream body while promoting various nutritional supplements. | Beneficial | Harmful | App (GYMKY)<br><br>Foods and food supplement (Prozis) | The influencer suggests that healthy nutrition is only possible with food supplements. The ideal of beauty portrayed (i.e. muscular, thin) is questionable and the influencer suggests that this ideal can only be achieved with his nutrition tips and the products he advertises. |
| Nutrition | Instagram: esmuellert<br><br>Follower: 12.800.000 | 307.782 | Three pictures showing the influencer preparing and eating peppers. The caption provides information on the nutritional value of peppers. | Beneficial | Beneficial | No ads | - |
| Nutrition | TikTok: markus_ruehl<br><br>Follower: 67.400 | 196.011 | The video shows the influencer sitting in a KFC restaurant while holding a KFC crispy chicken bucket. He promotes a KFC offer for the mass phase during training. | Harmful | Harmful | Fast food (Kentucky Fried Chicken) | The influencer promotes unhealthy foods (high in saturated fat and sugar) and advertises fast food products for training. |
| Nutrition | YouTube: Doctor Mike<br><br>Follower: | 192.700 | In the video the influencer provides information about potentially harmful contents in everyday foods (i.e. acids in rhubarb leaves) if they are unripe or wrongly prepared. | Neutral | Neutral | No ads | Questionable content since vegetables and fruits are portrayed in a harmful light and suggest that these foods should not be consumed. |

| | | | | | | | |
|---|---|---|---|---|---|---|---|
| Nutrition | Instagram: rezo<br><br>Follower: 1.750.000 | 162.806 | The picture shows the influencer eating fast food (kebab) and drinking sweetened beverages with a friend while sitting on the sidewalk. The caption to the post reads “best life”. | Harmful | Harmful | No ads | - |
| Nutrition | Instagram: teichners_pizza_palace<br><br>Follower: 181.000 | 160.703 | The video shows the influencer baking ciabatta with the recipe in the caption. | Beneficial | Neutral | No ads | - |
| Nutrition | Instagram: juliabeautx<br><br>Follower: 3.900.000 | 146.974 | Two pictures showing the influencer posing with donuts while promoting a christmas offer and lottery from a food producer (Desserts Factory). | Harmful | Harmful | Food delivery service (Flink)<br><br>Food producer (Desserts Factory) | Promotion of processed foods high in saturated fats and sugar. |
| Nutrition | Instagram: rezo<br><br>Follower: 1.750.000 | 141.672 | The picture shows the influencer cooking with a friend while the caption promotes a supermarket (Aldi) | Beneficial | Neutral | Supermarket (Aldi) | - |
| Reproductive Health | Instagram: copuleontour<br><br>Follower: 2.500.000 | 1.123.400 | The picture shows the influencer couple holding their baby. The caption explains that one of the influencers had a stroke and rehab during the past months and thanks her partner and the followers for their support. | Beneficial | Neutral | No ads | - |
| Reproductive Health | Instagram: anajohnson<br><br>Follower: 1.300.000 | 442.700 | Image and short video showing the moment the influencers receive the news that they are pregnant. | Beneficial | Neutral | No ads | - |
| Reproductive Health | Instagram: christian.wolf<br><br>Follower: 342.000 | 254.185 | Several pictures and videos of the influencers while they receive the news that they are pregnant and how they share this information with friends and family. | Neutral | Neutral | No ads | - |
| Reproductive Health | Instagram: danaemercer | 199.022 | Several pictures of the influencer in underwear while she is pregnant, showing her body | Beneficial | Beneficial | No ads | - |

| | | | | | | | |
|---|---|---|---|---|---|---|---|
| | Follower:<br>2.300.000 | | transformation (i.e. cellulite, stretch marks, hanging breasts). In addition, several pictures of instagram posts from other influencers regarding body positive content. The caption provides educational content about questionable beauty ideals on social media. | | | | |
| Reproductive Health | Instagram:<br>anajohnson<br><br>Follower:<br>1.300.000 | 140.605 | Several pictures of the influencer showing her way to get pregnant (i.e. fertility clinic, medication, food supplements). | Beneficial | Neutral | No ads | Although no ads or promotion of products, one picture shows her medication, including several food supplements. |
| Reproductive Health | Instagram:<br>fitnessoskar and healthy_mandy<br><br>Follower:<br>507.000<br><br>1.000.000 | 118.600 | The post includes one picture showing both influencers and one picture with a positive and negative pregnancy test. In the caption the influencers share their story of a miscarriage after the process of artificial fertilization. | Neutral | Neutral | No ads | - |
| Reproductive Health | Instagram:<br>danaemercer<br><br>Follower:<br>2.300.000 | 109.718 | In the picture the influencer compares pictures of herself from different angles while she is pregnant to point out questionable beauty ideals on social media. | Beneficial | Beneficial | No ads | - |
| Reproductive Health | Instagram:<br>danaemercer<br><br>Follower:<br>2.300.000 | 90.899 | Short video of the influencer as a postpartum mother informing about beauty ideals on social media compared to her body transformation during and after pregnancy (i.e. stretching mark, hanging breasts, belly fat). | Beneficial | Beneficial | No ads | - |
| Reproductive Health | Instagram:<br>danaemercer<br><br>Follower:<br>2.300.000 | 84.724 | Short video of the influencer as a postpartum mother informing about her body transformation during and after pregnancy (i.e. loose skin, cellulite, stretching mark, hanging breasts, belly fat). | Beneficial | Beneficial | No ads | |
| Reproductive Health | Instagram:<br>danaemercer<br><br>Follower:<br>2.300.000 | 78.440 | The post includes three pictures of the influencer in different poses to showcase the distorted display of beauty ideals on social media while promoting body positivity and educational content in the caption. | Beneficial | Beneficial | No ads | |

| | | | | | | | |
|---|---|---|---|---|---|---|---|
| Sexual Health | YouTube:<br>Noel Deyzel<br><br>Follower:<br>3.270.000 | 395.709 | Short video of the influencer posing in a tank top and flexing his muscles while talking about the effects of masturbation before working out. | Beneficial | Neutral | No ads | Promotion of questionable body image. |
| Sexual Health | Instagram:<br>jax.vita<br><br>Follower:<br>339.000 | 108.899 | The post is a short video with the influencer talking about the effects of taking and discontinuing birth control pills. | Beneficial | Neutral | No ads | - |
| Sexual Health | TikTok:<br>Gianna Bacio<br><br>Follower:<br>716.800 | 81.544 | The influencer is informing about intimate shaving, including tips to avoid skin irritation. | Beneficial | Neutral | Skincare product (Vagisan care) | - |
| Sexual Health | YouTube:<br>maiLab<br><br>Follower:<br>1.450.000 | 75.356 | In the video the influencer educates about sex and gender from a scientific and social perspective. | Beneficial | Beneficial | No ads | - |
| Sexual Health | YouTube:<br>Psych2Go<br><br>Follower:<br>10.800.000 | 58.565 | The video provides educational content about rare sexual disorders. | Beneficial | Beneficial | No ads | Potentially harmful for self-diagnosis of sexual disorders. |
| Sexual Health | YouTube:<br>Psych2Go<br><br>Follower:<br>10.800.000 | 51.052 | The video provides educational content about the effects of sex on the brain. | Beneficial | Beneficial | No ads | - |
| Sexual Health | YouTube:<br>Mama Doctor Jones<br><br>Follower:<br>1.270.000 | 26.374 | In the video the influencer who presents herself as gynecologist evaluates, criticizes, and debunks different statements from influencers regarding birth control pills. | Beneficial | Beneficial | No ads | - |
| Sexual Health | Instagram:<br>alexasearth &<br>elenacarriere | 17.830 | Several pictures of two influencers cuddling topless in bed. | Neutral | Neutral | No ads | - |

| | | | | | | | |
|---|---|---|---|---|---|---|---|
| | Follower:<br>200.000 & 460.000 | | | | | | |
| Sexual Health | YouTube:<br>Mama Doctor Jones<br><br>Follower:<br>1.270.000 | 18.399 | The influencer provides educational content on male birth control pills and debunks questionable information from another influencer (Johnny Harris). | Beneficial | Beneficial | No ads | - |
| Sexual Health | YouTube:<br>Mama Doctor Jones<br><br>Follower:<br>1.270.000 | 15.709 | In the video the influencer provides information on the safety of different birth control methods and possible (side)effects. | Beneficial | Beneficial | No ads | - |
| Sports, Training & Fitness | TikTok:<br>sydgrows<br><br>TikTok:<br>516.200 | 3.303.677 | In the video the influencer is shown talking to her friends in the gym instead of working out. | Neutral | Neutral | Clothing (Gymshark) | - |
| Sports, Training & Fitness | YouTube:<br>Fabio Wimber<br><br>Follower:<br>7.460.000 | 2.746.514 | The influencer is shown biking through a city while performing different Stunts. In one sequence he consumes and promotes an energy drink. | Beneficial | Harmful | Energy drink (Red Bull)<br><br>Clothing (POC) | The video promotes sweetened beverages and connects athleticism with the consumption of energy drinks. |
| Sports, Training & Fitness | Instagram:<br>cbum<br><br>Follower:<br>17.000.000 | 2.490.448 | The post contains a picture of the influencers legs in shorts while flexing his muscles and a video performing a workout. | Neutral | Harmful | No ads | Promotion of questionable body type (i.e. muscular). |
| Sports, Training & Fitness | YouTube:<br>Fabio Wimber<br><br>Follower:<br>7.460.000 | 2.025.129 | The video shows the influencer biking through the city and forest performing different stunts. | Neutral | Neutral | Clothing (Sick Series) | - |
| Sports, Training & Fitness | Instagram:<br>cbum<br><br>Follower:<br>17.000.000 | 1.850.025 | Several pictures of the influencer preparing and performing in a bodybuilding show. | Neutral | Harmful | No ads | Promotion of questionable body image (i.e. muscle mass). |

| | | | | | | | |
|---|---|---|---|---|---|---|---|
| Sports, Training & Fitness | Instagram: cbum<br><br>Follower: 17.000.000 | 1.776.346 | The video shows the influencer working out in a gym. | Neutral | Harmful | No ads | Promotion of questionable body image (i.e. muscular) |
| Sports, Training & Fitness | YouTube: Lilly Sabri<br><br>Follower: 4.500.000 | 1.652.597 | In the video the influencer is shown working out while promoting her own workout app. Furthermore, she promises in the video and caption to loose belly fat within 7 days if one follows her workout routine. In the caption the cookbook of the influencer is promoted promising to help against inflammation and increasing immunity. | Beneficial | Harmful | App (leanwithlilly)<br><br>Cookbook (leanwithlilly)<br><br>Food supplements (Optimum Nutrition) | Promotion of questionable body type (i.e. thin, muscular). The video suggests that one should work out to lose weight and not other health benefits. Misleading promises that visible physical changes will occur within seven days.<br>Promotion of food supplements. |
| Sports, Training & Fitness | YouTube: Fabio Wimber<br><br>Follower: 7.460.000 | 1.532.111 | The video shows the influencer biking through the city performing different stunts. | Beneficial | Harmful | Energy drink (Red Bull)<br><br>Clothing (Sick Series) | Promotion of sweetened beverages (Red Bull). |
| Sports, Training & Fitness | TikTok: leanadeep<br><br>Follower: 12.400.000 | 936.215 | In the video the influencer provides tips for workouts to increase performance and mobility. | Beneficial | Beneficial | Clothing (gymshark) | - |
| Sports, Training & Fitness | Instagram: cbum<br><br>Follower: 17.000.000 | 934.046 | The post contains several pictures of the influencer posing in a gym while promoting his own product line of clothing. | Neutral | Neutral | Clothing (cbumfitness) | - |
